# ICD-10 Code Ambiguity Obscures Treatment-Eligible Adults with Spinal Muscular Atrophy: A Single-Center Chart Review and Patient Outreach Study

**DOI:** 10.64898/2026.06.07.26355122

**Authors:** Gabriel Holly, Bryce Bean, Haidy Beshay, Gabrielle Edwards, Nicholas Streicher

## Abstract

**Background:** Three disease-modifying therapies (DMTs) for spinal muscular atrophy (SMA) have been approved since 2016, yet many adults remain untreated. Identifying them depends on ICD-10 codes that capture SMA but do not reliably distinguish it from other related conditions. We examined, in one U.S. health system, both patients’ engagement with therapy and the accuracy of the codes used to find them.

**Methods:** We conducted a retrospective chart review of adults in an academic health system identified by SMA-associated ICD-10 codes, with manual adjudication of diagnosis and DMT status. Confirmed SMA-positive, DMT-naïve patients were invited to a structured telephone interview on treatment awareness and barriers.

**Results:** Of 60 charts, 22 (36.7%; 95% CI 25.6–49.3%) were appropriately coded for SMA or a related disorder; only 16 (26.7%) had molecularly confirmed SMA. The other 38 (63.3%) were miscoded, spanning spinal and bulbar muscular atrophy, asymptomatic carriers, prenatal screening, and conditions unrelated to SMA. Ten of the 16 confirmed patients (62.5%) were DMT-naïve; one was interviewed, one declined, and eight could not be reached. The non-response is itself a finding: the patients least visible to administrative data are the hardest to reach.

**Conclusions:** ICD-10 ambiguity is a barrier to treatment access in adult SMA, as is loss to follow-up. We make two recommendations: continuous documentation-coding alignment that uses natural language processing to verify the genetic precondition, and type-specific SMA codes (subcodes for Types 0–4) anchored on molecular SMN1 confirmation. Together these would support cohort identification, outreach, and evidence generation without adding to clinician burden.

## Background

Spinal muscular atrophy (SMA) is an autosomal recessive neurodegenerative disorder caused by biallelic loss-of-function mutations in the survival motor neuron 1 (SMN1) gene on chromosome 5q13; clinical severity is graded from Type 0 to Type 4, with more copies of the related SMN2 gene producing milder disease [1]. U.S. newborn screening places birth prevalence near 1 in 14,694 [2]; adults now make up roughly half of the prevalent SMA population, an estimated 8,000 to 10,000 individuals [3,4].

Once untreatable, SMA now has three disease-modifying therapies (DMTs): nusinersen (2016) and risdiplam (2020), both labeled for pediatric and adult patients, and onasemnogene abeparvovec, a gene therapy approved in 2019 as an intravenous infusion for children under two and in November 2025 as an intrathecal formulation for ages two and older. Real-world evidence supports adult efficacy [5,6], yet 21% of Type 3 patients (predominantly adults) remained untreated in a 2023 Italian nationwide cohort even within a publicly funded system [7].

Most adults with SMA today reached adulthood before any DMT existed, often diagnosed before molecular testing was routine. Those with milder Type 3 or Type 4 disease, who may have had little reason to stay in specialist care once stable, are therefore among the least likely to be in active neuromuscular care or reliably ascertained from health-system data.

Finding these patients means searching coded health-system data, where the primary SMA code is an unreliable filter, conflating true SMA with similarly named conditions such as spinal and bulbar muscular atrophy (SBMA) [8]. The problem is recognized but unmeasured. In a large claims-based SMA cohort, Belter and colleagues noted that these codes neither distinguish SMA subtypes nor reliably identify true cases, yet did not quantify the error against chart review [9]. SMA is not unique: only 56.4% of monogenic epilepsy patients receive a syndrome-specific code when one exists [10], and codes match the clinical note far less often when the code is entered apart from the note [11].

We therefore examined adults with SMA in a single U.S. health system on two fronts: their awareness of and engagement with DMT, and the accuracy of the ICD-10 codes used to identify them.

## Methods

### Study Design and Setting

We conducted a single-center retrospective chart review with an embedded exploratory patient-interview component within an academic multi-hospital health system in the Mid-Atlantic United States. The study was approved by the MedStar Health Research Institute Institutional Review Board (Protocol STUDY00008773); interview participants provided verbal informed consent documented in study records, and a HIPAA waiver was granted for the chart review.

### Cohort Identification

On October 29, 2025, we queried the health-system electronic medical record (EMR) for all adults (≥18 years) with any inpatient or outpatient encounter over the preceding 10 years coded with an SMA-associated ICD-10-CM code: G12.0 (infantile SMA type I), G12.1 (other inherited SMA), G12.8 (other SMA and related syndromes), or G12.9 (SMA unspecified). Charts were first reviewed for patients seen in the institutional Muscular Dystrophy Association (MDA) clinic, then for patients seen elsewhere within the system.

### Chart Adjudication

Charts were reviewed initially by the study team and adjudicated by NS to determine: (i) presence or absence of confirmed SMA based on documented SMN1 deletion testing, geneticist or neurologist diagnostic statement, or both; (ii) the actual clinical condition documented if not SMA; (iii) DMT status (current, prior, or no documented therapy); and (iv) the specific ICD-10 code(s) responsible for cohort inclusion.

Charts were classified as: (a) confirmed SMA, appropriately coded; (b) other appropriately coded motor neuron condition (e.g., Hirayama disease coded as G12.8); (c) SBMA miscoded as SMA; (d) asymptomatic SMA gene carrier identified via family or carrier screening; (e) prenatal carrier screening case; (f) no relevant neuromuscular diagnosis identified; or (g) unrelated diagnosis with SMA-related code applied in error.

## Interview Component

Patients with confirmed SMA but no documented DMT were invited to the interview, which aimed to distinguish among scenarios not separable from administrative data: unawareness of therapy, awareness with insurance or logistical barriers, relocation with or without continuity of care, or active decline.

Eligible patients were contacted by telephone using EMR-listed information, with up to three attempts at varied times of day. Consenting participants completed a structured interview covering five domains: (1) timing and circumstances of SMA diagnosis; (2) awareness of currently available DMTs; (3) prior provider conversations about treatment; (4) perceived barriers to treatment initiation; and (5) experience navigating adult neuromuscular care. The full instrument is provided as Additional file 1.

### Data Analysis

The proportion of code-flagged patients with adjudicated SMA was calculated as a positive predictive value (PPV), with 95% confidence intervals (CI) computed using the Wilson score method. Code-specific patterns are reported as counts and proportions stratified by ICD-10 code. Analyses were performed in IBM SPSS Statistics. Interview content is reported as descriptive narrative.

## Results

### Cohort Composition and Adjudication

Sixty unique adult charts met criteria for review (Figure 1). Only 16 had confirmed SMA, a positive predictive value of 26.7% for the combined code set (16/60; 95% CI 17.1–39.0%); counting six other motor neuron conditions correctly captured (five Hirayama disease, one distal SMA), the value for any code-appropriate diagnosis rose to 36.7% (22/60; 95% CI 25.6–49.3%). The remaining 38 charts (63.3%) were miscoded.

**Figure 1.**
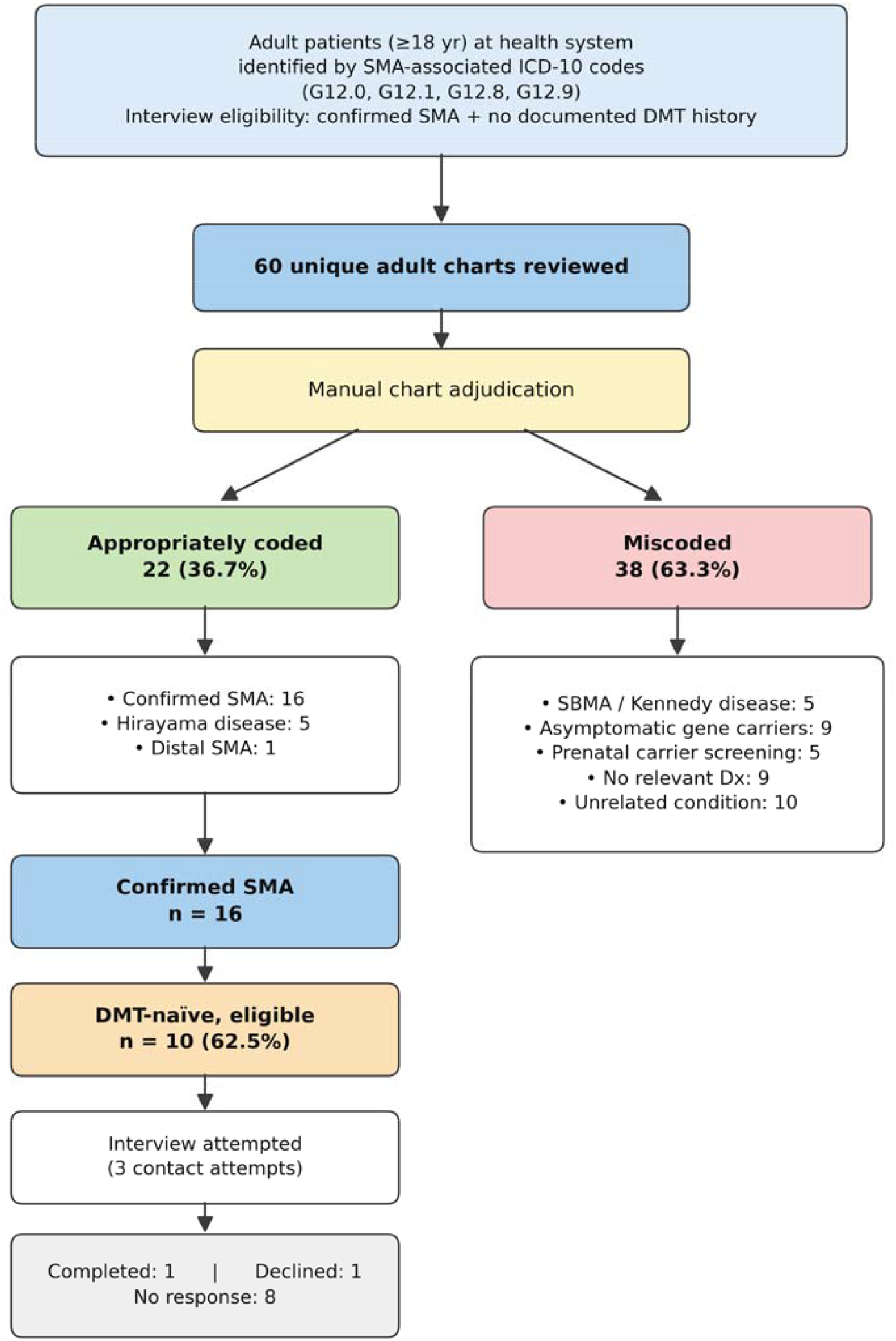
Patient flow.

### Patterns of Miscoding

The 38 miscoded charts fell into five categories (Table 1, Figure 2): five (8.3%) had SBMA miscoded as SMA; nine (15.0%) were asymptomatic gene carriers found through family or partner screening; five (8.3%) entered through prenatal carrier screening; nine (15.0%) carried an SMA-related code with no documented neuromuscular diagnosis; and ten (16.7%) had wholly unrelated conditions, such as Huntington disease, multiple system atrophy, and muscular dystrophy, often from a code added during early workup and never removed.

**Table 1.**
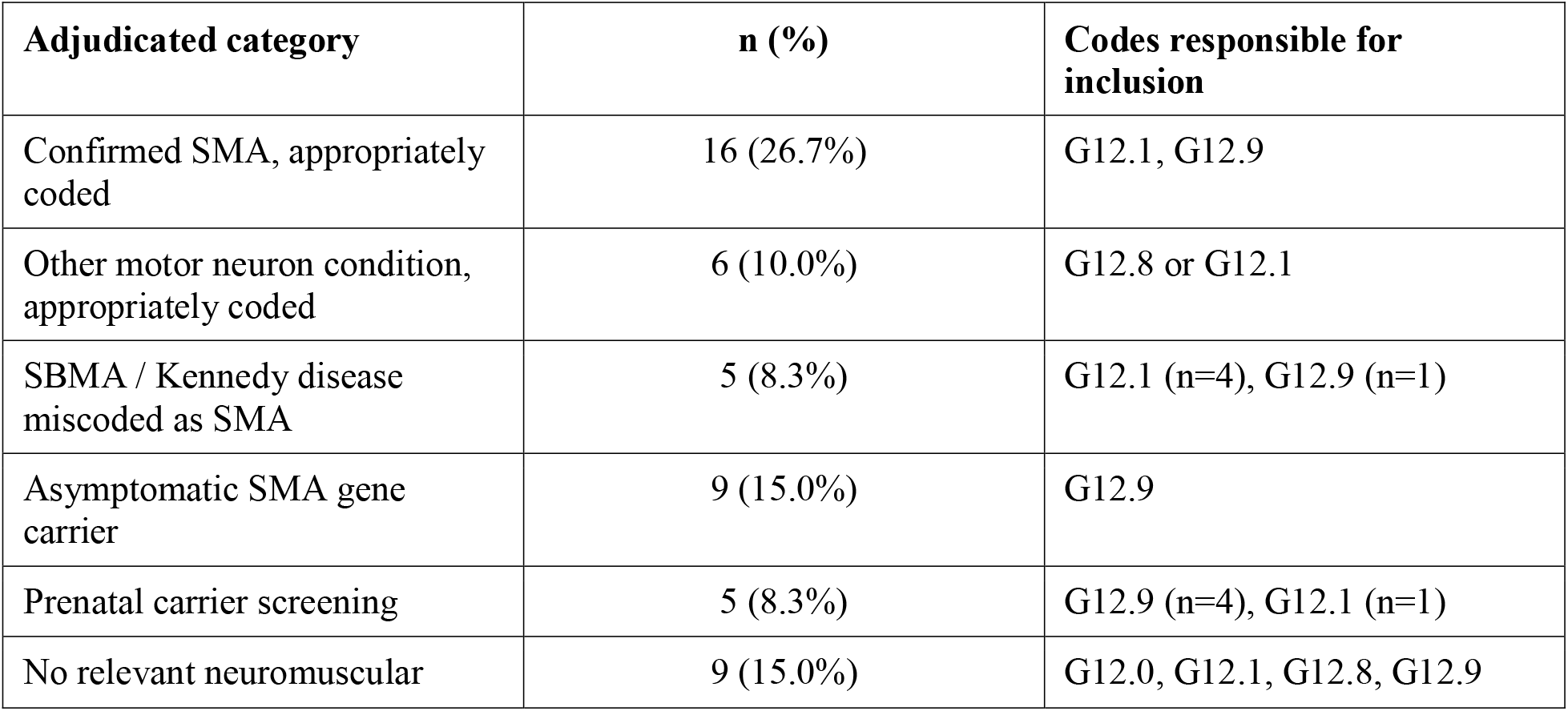
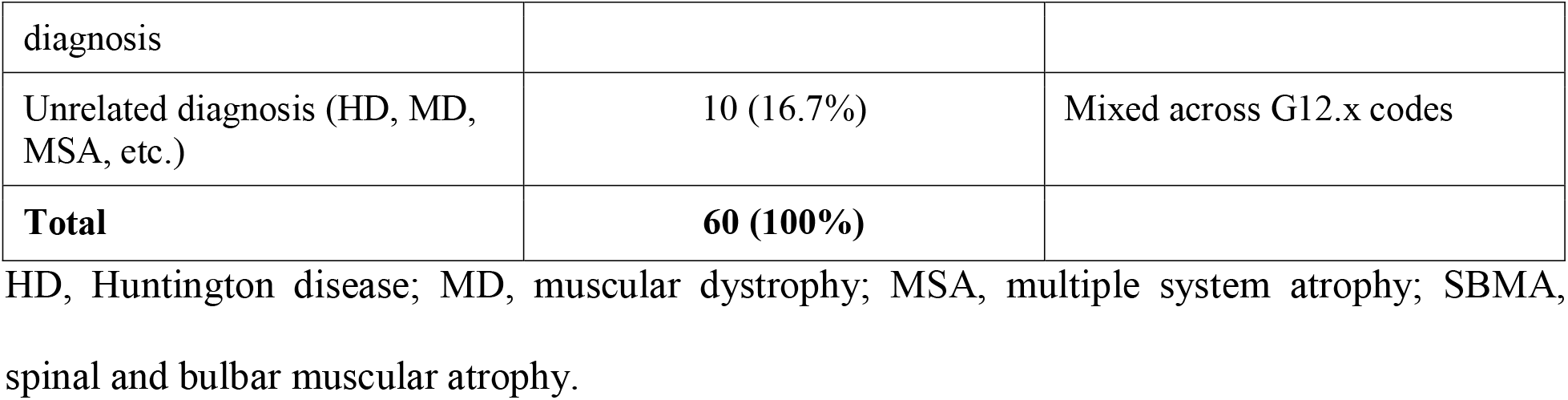
Distribution of patients by adjudicated diagnosis and ICD-10 code.

**Figure 2.**
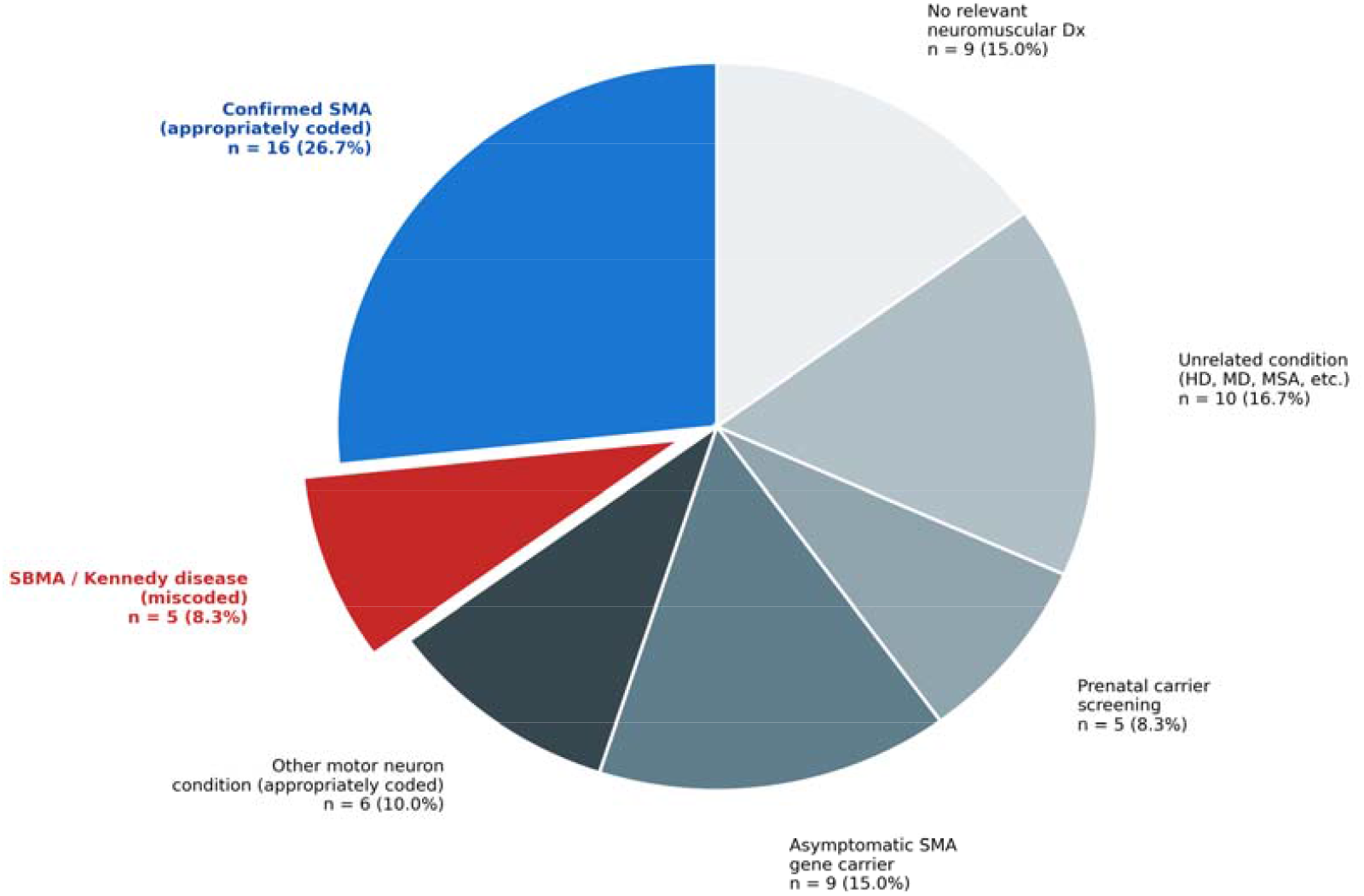
Adjudicated diagnoses.

### Treatment-Eligible Cohort, Contact Outcomes, and Interview Findings

Of the 16 confirmed SMA patients, five had current or prior DMT documented and one had died, leaving ten SMA-positive, DMT-naïve patients eligible for interview, or 62.5% of confirmed cases. Of these ten, one completed a structured interview, one declined, and eight (80%) could not be reached after three attempts.

The participant had Type 3 SMA diagnosed before the treatment era, reported limited awareness that DMT is now available to adults, and cited insurance as a likely barrier, but expressed interest in re-engaging.

## Discussion

SMA-associated codes are accurate less often than other neurological codes, correct about two-thirds of the time for amyotrophic lateral sclerosis [12] and about half for muscular dystrophy [13]. Of the 60 adults flagged, fewer than one in three had confirmed SMA, with the remainder including related but alternative diseases, conditions properly coded but genetically distinct, prenatal screening, and others altogether miscoded. This error was long suspected but never quantified against charts in SMA until this study [9].

When patients are miscoded, the true disease cohort and its prevalence cannot be reliably established. Coding is the foundation of most disease queries, so inconsistent recording undermines efforts to study prevalence, treatment patterns, and natural history, from a single system to larger national or international disease registries. Correcting this requires aligning codes with the clinical record and adding new SMA codes anchored on molecular confirmation. Alignment ensures codes are applied as documented, and the molecular anchor ensures the codes themselves are precise.

The one patient we interviewed, an eligible but untreated adult, had fallen out of specialist care and did not know disease-modifying therapy now existed, and once aware pointed to insurance coverage as a further barrier. A single interview is illustrative rather than representative, and it puts a human face on the data. Its account is consistent with awareness and access barriers reported in larger SMA cohorts [7]. The patients hardest to find are also the hardest to re-engage.

### Implications for Practice and Research

Although SBMA (five charts, 8.3%) [8] and distal SMA (one chart) can resemble SMA clinically, they are distinct genetic diseases that may be under-recognized [14] yet coded as SMA, while true SMA can be missed [15]. Rare-disease coding reform is already underway elsewhere. Europe developed a dedicated rare-disease coding system, Orphacodes, to capture diseases that ICD-10 cannot distinguish [16]. In the United States, the 2023 leukodystrophy effort [17] offers a model, advanced through the National Center for Health Statistics (NCHS) process: require SMN1 confirmation for the main SMA code, add subcodes for Types 0–4, and retire the “other” and “unspecified” categories that lump together conditions now considered distinct entities.

Codes and documentation must be kept aligned continuously, using natural language processing and structured clinical data. This alignment should occur at note signing, during regular sweeps of code-flagged patients, and before research analysis [18]. Early evidence supports this approach [19,20,21], but it must be built carefully to avoid introducing new data-integrity issues. General-purpose language models should not be used on their own, as their performance without proper training is unreliable [22,23]. These models should be specially trained rather than generic and should assist human reviewers in maintaining data integrity rather than replacing them, as liminal cases often require expert decision-making.

Rare-disease codes left on the problem list after workup should be removed once follow-up no longer supports them, which could aid clinical practice and research. Data integrity extends beyond accurate codes to complete records across systems. A single institution cannot tell loss to follow-up from care delivered elsewhere, so privacy-preserving record linkage could separate attrition from transfer for both treatment and diagnostic purposes.

### Limitations

We identified our cohort using the same codes we found to be unreliable, so we may have missed true cases coded differently or miscoded. SMA is clinically distinctive, and a code is typically entered at some point during patient contact, so the harder problem is separating true cases from the miscoded ones. For cases we could not reach or that did not answer our calls, we cannot confirm treatment elsewhere. Our single completed interview cannot support a thematic analysis, and a single-center rate may not transfer to other coding environments, although the current literature suggests this pattern holds in claims databases and elsewhere.

## Conclusions

Diagnostic miscoding is a barrier to treatment access in adult SMA, as is loss to follow-up. Many adults lost contact with the medical system before treatments emerged later in their disease course. Fewer than one in three SMA-coded patients had confirmed disease, and 80% of treatment-eligible adults could not be reached.

We propose continuous documentation-coding alignment paired with type-specific SMA codes; together these would support cohort identification, outreach, and evidence generation without adding to clinician burden. This model in fact generalizes to other genetic diseases, especially those where clinical knowledge has outpaced the codes and novel treatments emerge in adulthood. Research and care increasingly depend on large-scale data and the models built on it, so accurate coding matters more than ever; analyses can only reflect the diseases as they were recorded. This returns coding to its original purpose, before billing claimed it: to faithfully represent what the clinician documented.

## Supporting information

Supplemental Interview

## Data Availability

The data supporting the findings of this study contain protected health information and are not publicly available. De-identified data may be made available from the corresponding author on reasonable request, subject to institutional and privacy approvals.

## Declarations

### Ethics approval and consent to participate

This study was approved by the MedStar Health Research Institute Institutional Review Board (Protocol STUDY00008773) and was conducted in accordance with the Declaration of Helsinki. A HIPAA waiver was granted for the retrospective chart review. All interview participants provided verbal informed consent before the interview, documented in study records.

### Consent for publication

Not applicable. The single interview narrative is described in aggregate terms with no identifying personal details.

### Availability of data and materials

De-identified chart review data and interview notes are not publicly deposited because of the small adult SMA cohort at a single center and the consequent risk of patient re-identification. Aggregate data summaries are available from the corresponding author on reasonable request, subject to MedStar Health Research Institute IRB approval. The interview instrument is provided as Additional file 1.

### Authors’ contributions

NS conceived the study, designed the protocol, and supervised the research. GH led chart adjudication and patient outreach, and drafted the manuscript. BB, HB, and GE contributed to chart review and patient outreach coordination. NS and GH analyzed the data and produced the figures. All authors contributed to interpretation of findings, manuscript review, and approved the final version.

## Acknowledgements

The authors thank the institutional Department of Neurology for support, and the patients and families who participated in this study.

## Additional files

Additional file 1 (.docx): Patient Interview Instrument. The structured telephone interview administered to confirmed SMA patients without documented disease-modifying therapy.

## Abbreviations

SMA: spinal muscular atrophy
SMN1: survival motor neuron 1 gene
DMT: disease-modifying therapy
ICD-10-CM: International Classification of Diseases, Tenth Revision, Clinical Modification
SBMA: spinal and bulbar muscular atrophy
PPV: positive predictive value
EMR: electronic medical record
MDA: Muscular Dystrophy Association
NCHS: National Center for Health Statistics
CI: confidence interval.

