## Supplemental Interview for "ICD-10 Code Ambiguity Obscures Treatment-Eligible Adults with Spinal Muscular Atrophy: A Single-Center Chart Review and Patient Outreach Study"

**Additional file 1: Patient Interview Instrument**

Protocol title: Understanding the Diagnostic Pathway and Treatment Experience of Patients with Spinal Muscular Atrophy (SMA).

The following structured telephone interview was administered to confirmed SMA patients without documented disease-modifying therapy. Interviews were conducted by study investigators; eligible patients were contacted at multiple times of day, with up to three attempts.

**Diagnostic Journey**

When were you diagnosed with spinal muscular atrophy (SMA)?

When did you first notice symptoms related to muscle weakness?

How affected are you now? (none / minimal / moderate / severe)

How long did it take to receive a diagnosis of SMA, and how many doctors did you see during this process?

Have you experienced symptoms related to muscle atrophy, motor delays, or respiratory issues? If yes, when did these first appear?

Have you ever experienced issues with hand grip or fine motor skills? If so, when did these symptoms start?

Were you referred to a neurologist?

Have you had an EMG? An MRI? Any other diagnostic tests related to SMA?

Have you undergone genetic testing for SMA?

Do any family members have genetic testing for SMA or related conditions?

Have you seen a genetic counselor regarding SMA?

**Treatment**

Have you previously been on a disease-modifying treatment for SMA? (yes / no)

***If previously treated:***

How old were you when you started, and what were the treatment dates for each drug (nusinersen [Spinraza]; risdiplam [Evrysdi])?

Why did you stop taking the medication?

***If not currently treated:***

Is there a particular reason you are not on any medication? (probe as necessary with the questions below)

Do you have minimal symptoms and feel treatment is unnecessary?

Are you unaware of available treatments?

Are you interested in treatment but face difficulties in accessing it? If so, what difficulties are you facing?

**Demographics**

What is your educational status?

Are you employed or studying? What do you do?

Do you have a caregiver, or someone who helps with your care? Do you live alone?

Do you have reliable transportation for medical appointments and treatment?

How many times have you been to the emergency room in the past 12 months?

Are you insured? What is your insurance?

**Closing**

That concludes our survey. Before I hang up, what is the best email to reach you to send the gift card? Also, are you willing to be contacted about other studies?
